# COVID-19 Related Chemosensory Changes in Individuals with Self-Reported Obesity

**DOI:** 10.1101/2021.02.28.21252536

**Authors:** Surabhi Bhutani, Géraldine Coppin, Maria Geraldine Veldhuizen, Valentina Parma, Paule Valery Joseph

## Abstract

**Background/objectives:** Individuals with obesity show alterations in smell and taste abilities. Smell and taste loss are also the most prominent neurological symptoms of COVID-19, yet how chemosensory ability present in individuals with obesity with a positive COVID-19 diagnosis is unknown.

**Subjects/Methods:** In this secondary analysis of a cross-sectional global dataset, we compared self-reported chemosensory ability in participants with a respiratory illness reporting a positive (C19+; n = 5156) or a negative (C19−; n = 659) COVID-19 laboratory test outcome, who also self-reported to be obese (C19+; n = 433, C19−; n = 86) or non-obese.

**Results:** Compared to the C19− group, C19+ exhibited a greater decline in smell, taste, and chemesthesis during illness, though these symptoms did not differ between participants with obesity and without obesity. In 68% of participants who reported recovery from respiratory illness symptoms (n=3431 C19+ and n= 539 C19−), post-recovery chemosensory perception did not differ in C19+ and C19− diagnosis, and by self-reported obesity. Finally, we found that all chemosensory and other symptoms combined predicted the C19+ diagnosis in participants with obesity with a moderately good estimate (63% accuracy). However, in C19+ participants with obesity, we observed a greater relative prevalence of non-chemosensory symptoms, including respiratory as respiratory and GI symptoms.

**Conclusions:** We conclude that despite a presumed lower sensitivity to chemosensory stimuli, COVID-19 respondents with obesity experience a similar self-reported chemosensory loss as those without obesity, and in both groups self-reported chemosensory symptoms are similarly predictive of COVID-19.

## INTRODUCTION

According to the World Health Organization, globally 13% of adults aged 18 years and over reported to have obesity in 2016 (1). Within the context of the ongoing COVID-19 pandemic, intriguingly, countries with the highest prevalence of obesity also recorded a high death rate from COVID-19 infection (2). Although, an increased in susceptibility to viral infection with obesity is unknown, a recent review by Stefan et al. concluded that obesity is a strong and independent determinant of morbidity and mortality in patients infected with SARS-CoV-2, the virus responsible for COVID-19 infection (3). A recent analysis also indicated that COVID-19 mortality in patients with obesity is higher than that of other comorbidities, including diabetes, hypertension, asthma, and cancer (4). In addition to greater risk for COVID-19 related poor health outcomes (3,5,6), patients with obesity are more likely to require hospitalization, especially in young adults with a Body Mass Index (BMI) >30 kg/m (7). Overall, current evidence suggests that obesity significantly interacts with the pathogenesis of COVID-19. Despite this risk, COVID-19 chemosensory symptoms have not yet been systematically assessed in this population group.

Disturbances in smell and taste emerged as a predominant neurological symptom of COVID-19 infection, with 77 percent of COVID-19 patients reporting sudden olfactory and gustatory dysfunctions in a recent meta-analysis (8). In a recent analysis, we also reported quantified smell loss as the best predictor of COVID-19, compared to other common non-chemosensory symptoms (9). However, these studies did not delineate COVID-19-related chemosensory impact in patients with obesity. This is especially important because individuals with obesity typically have existing lower taste sensitivity, and lower capacity to detect and identify odors than individuals without obesity (10). Particularly, excessive body weight has been shown to be associated with impaired taste for sweet and salty foods, alteration in fat/fatty acid-sensing, reduced ability to identify correct taste (11–13) in taste detection thresholds (11–15). These obesity-related chemosensory dysfunctions are driven by production of pro-inflammatory factors from adipose tissues, leading to impairment in olfactory receptors (16) and a decline in taste bud and taste progenitor cells (17,18), respectively. Considering that marked inflammation with obesity also seems to favor viral infections (19,20), the interaction between existing chemosensory deficiency in adults with obesity and COVID-19 related chemosensory impairments are unknown.

Existing gustatory and olfactory sensory deficiency due to obesity may mask the viral-induced diminished taste and smell self-reported experiences, leading to a higher portion of undetected cases in this population (21), we need a better understanding of how chemosensory profile changes in patients with obesity. Furthermore, in light of the potential for using oro-naso sensory perception as an early marker of SARS-CoV-2 infection (22–24), it needs to be assessed whether the predictive relation between the chemosensory loss and COVID-19 illness generalizes to participants with obesity. With continually increasing death rates projected well into 2021 using a second statistical model (25), and a wave of infections sweeping through countries worldwide, it is imperative to understand the impact of SARS-COV-2 virus on chemosensory dysfunction in COVID-19 patients in the high-risk category such as populations with excessive body weight. Here, we systematically describe and compare the chemosensory perception (smell, taste, and chemesthesis) and related symptomatology in COVID-19 in non-hospitalized adults with or without self-reported obesity. Following our pre-registered analysis plan (26), we hypothesized that the participants with obesity will report less smell loss during COVID-19 illness. We also hypothesize that smell loss will be less predictive of COVID-19 diagnosis than the participants without obesity.

## METHODS

### Study Design

To the best of our knowledge, this study is the first to assess chemosensory alterations in adults with obesity and COVID-19. We conducted a secondary analysis of cross-sectional survey data collected between April 7th and November 4th, 2020 using the Global Consortium for Chemosensory Research (GCCR) core questionnaire. This crowdsourced survey collected data from community-dwelling individuals via social and traditional media as well as the GCCR website. It was also presented to clinicians to relay to their patients. This survey, currently deployed in 32 languages, used binary-response and categorical questions, as well as visual analog scales to measure self-reported chemosensory ability and other symptoms in adults with ongoing or recent respiratory illnesses (22). We also collected self-reported data on the presence of pre-existing diseases, including our condition of interest, obesity, as well as other COVID-19 symptoms All participants included in the study were: 1) ≥18 years old, 2) had a (suspected) respiratory illness within the past two weeks, 3) had onset of respiratory illness after January 1, 2020, 4) reported COVID-19 diagnosis via laboratory test (viral PCR or antigen test). Respondents who did not report having any illness or symptoms within the last two weeks, who had multiple responses, or who responded “Don’t know” or “Other” when asked about their diagnosis of COVID-19, were excluded from the analyses. To investigate the recovery of chemosensory functions, only participants who reported the date of onset of respiratory illness symptoms were included. The original study was approved by the Office of Research Protections of The Pennsylvania State University (STUDY00014904). The hypotheses and analyses in this manuscript were pre-registered at https://osf.io/xf25v (26) and the research compendium including data files and analysis scripts is available at https://osf.io/rbcty/. Specifically, our analyses aimed to describe chemosensory perception and related symptomatology during the COVID-19 illness (Aim 1) and post-vs pre-COVID-19 diagnosis (Aim 2), in participants with self-reported obesity vs without obesity. We predicted lower ratings for smell, taste, and chemesthesis, and more severe COVID-19 symptoms in participants with obesity, compared to without obesity. We also speculated smaller differences in ratings for smell, taste, and chemesthesis perception post-vs pre-COVID diagnosis in participants with self-reported obesity. Post-COVID-19 chemosensory recovery (Aim 3) was also tested, hypothesizing that participants will have lower ratings for smell, taste, and chemesthesis post-COVID-19 recovery in participants with self-reported obesity vs without obesity. Additionally, we assessed COVID-19 severity as measured based on the sum of reported symptoms (Aim 4), and the ability of smell ratings to predict COVID-19 diagnosis (Aim 5), in participants with self-reported obesity vs without obesity.

A departure from the pre-registered analyses is the inclusion of age as a factor in all analyses, following differences in age we observed between groups. We also report the unregistered analysis of pre-illness ratings, an important addition given the previously reported decreased sensitivity for participants with obesity compared to those without obesity.

### Participant Description

A convenience sample of 52334 volunteers accessed the GCCR questionnaire. Of those individuals, 5815 met the inclusion and exclusion eligibility criteria and were included in the final analysis. A positive COVID-19 diagnosis (C19+) was determined using the self-reported data from COVID-19 lab test or clinical exam outcome. All C19+ patients were further categorized into having obesity if they reported it as one of the pre-existing disease conditions in the questionnaire. C19+ patients who did not report having any medical condition or did not answer this question on pre-existing disease conditions were categorized as controls without obesity. We also included a control group of participants without obesity. See Figure 1. for a flow diagram of the inclusion of participants into the various groups.

**Figure 1.**
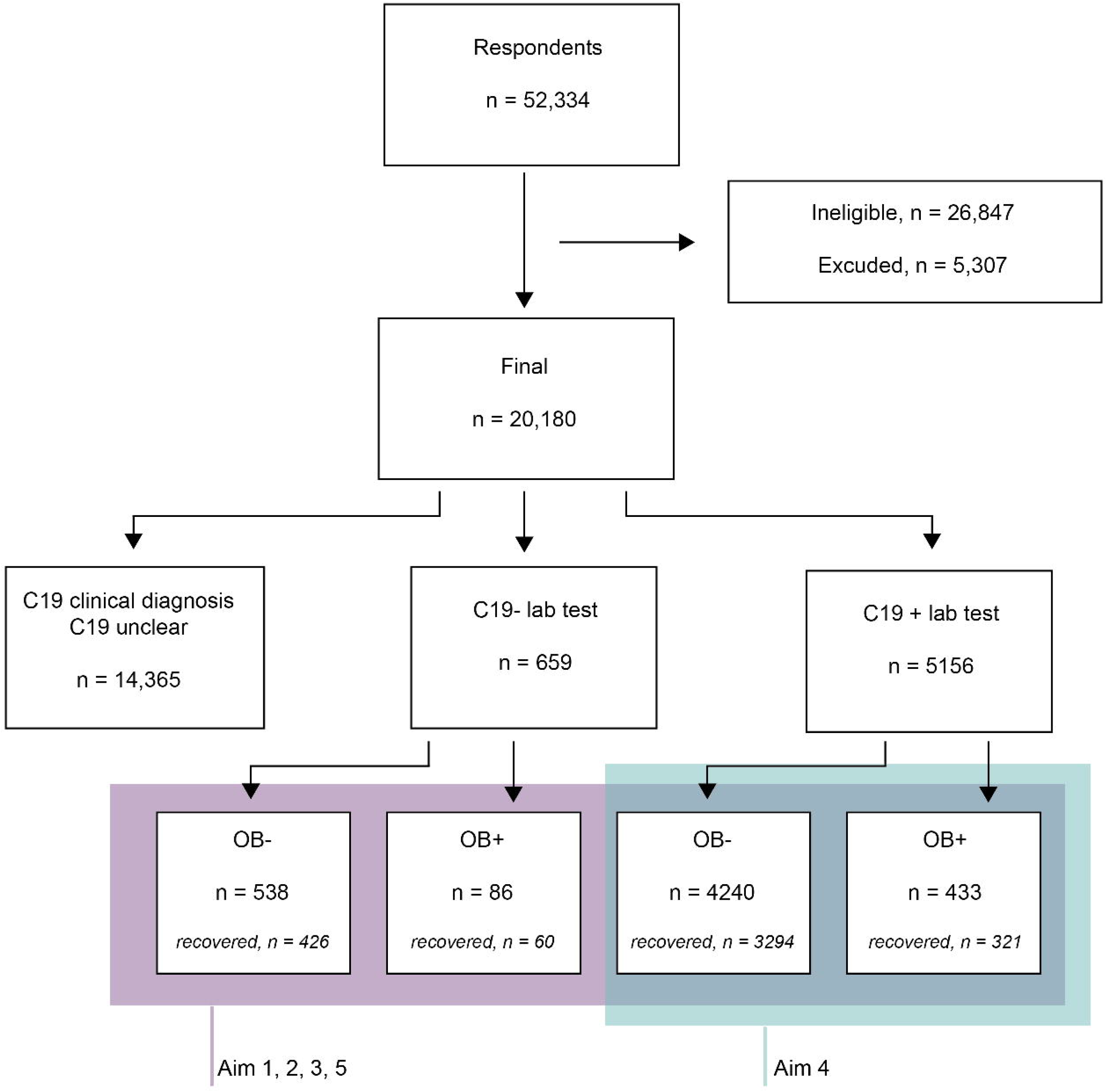
Flow Diagram of Study Participants Based on the STrengthening the Reporting of OBservational Studies in Epidemiology (STROBE) guidelines. Participants included in the prediction of COVID-19 status in participants with obesity vs without obesity are framed in blue. Participants framed in purple are included in all other analyses. n = number of participants; OB+ = self-reported presence of obesity; OB− = self-reported presence of obesity; COVID diagnosis unclear = responses “No - I do not have any symptoms”, “Don’t know” or “Other” to survey Question 8 (“Have you been diagnosed with COVID-19?”).

### Statistical Analyses

Statistical analyses were conducted in R (27) via RStudio. The annotated scripts, the information on the computational environment, and dependencies shared for future reproducibility will be found, upon acceptance of the manuscript, at the OSF project link, which includes directions to the GitHub page at which the code is stored.

No negative value appeared in the survey responses. Whenever in question 38 (prior conditions), no response was provided or the option “None” was checked, the response was imputed as indicating no prior conditions. Prediction targets were never imputed. All open-ended questions were excluded as they are incompatible with model generalization.

### Demographics

Cognizant of possible null effects in all our analyses, we opted to implement a Bayesian approach, which allows us to estimate the strength of the evidence supporting the null hypothesis. To test via a between-participant sequential Bayes factor design whether a difference between groups was present (H1) or absent (H0), we conducted Bayesian linear regressions with the lmBF function from the BayesFactor package (28). We used the default Cauchy prior on the effect sizes under the H1 as the scale parameter spread, which was set at its default value of r = sqrt (2)/2. To test for a difference in age between groups, we used the following full model: Age ~ COVID diagnosis + Obesity Age + COVID diagnosis x Obesity. Additive models (no interaction) and main effect models were also computed and compared to determine the model that best explained the data pattern, aka the model comparison with the most extreme Bayes Factor. Please refer to **Supplementary Table 1** for the inference rules, which follows the classification scheme proposed by Lee and Wagenmakers (29) and adjusted from (30). To interpret the strength and the direction of the effects identified, we have additionally sampled from the models’ posterior distributions (iterations = 1e4). To test for gender differences between groups, we calculated probability tables of women and men in each of the COVID-19 and obesity groups and tested for distribution differences with Pearson’s chi-square tests with the R base function “prop.test”. We used an alpha of 0.05 to determine significance.

### Self-reported Chemosensory perception analyses

For chemosensory perception analyses, we also conducted Bayesian linear regressions with the lmBF function. The full model included the following terms: Dependent variable ~ COVID diagnosis + Obesity Age + COVID diagnosis x Obesity. Additive models (no interaction) and main effect models were also computed and compared to determine the model that best explained the data pattern. Age was included in all models to factor in significant associations between age and obesity. We used “before illness”, “during illness”, “change due to illness” (“before illness” minus “during illness”) and “recovery” (“after illness” minus “during illness”) separately as dependent variables.

### Other illness symptomatology analyses

To assess whether participants with obesity experience more and/or different symptoms from those without obesity, we summed all symptoms that participants reported (each symptom that was reported was assigned a value of 1). We then conducted Bayesian linear regressions with the lmBF function as above with summed symptoms as the dependent variable (as above in the chemosensory analyses). We operationalized disease duration as the number of days since onset of the illness and used “days since onset” as the dependent variable in Bayesian linear regression (models as above). For the subset of COVID-19 patients only, we calculated probability tables for the likelihood of experiencing a given symptom for the participants with and without obesity and tested for distribution differences with chi-square tests (details as above under demographics). We used an alpha of 0.05 to determine significance.

### Model accuracy for predicting COVID-19 illness

To deal with binary classification problems in the presence of imbalanced classes, we used the ROSE (Random Over-Sampling Examples) package (31), which generates synthetic balanced samples and thus allows to strengthen the subsequent estimation of any binary classifier. To measure model quality, receiver operating characteristic (ROC) were visualized via the pROC package (32) based on the calculation of hold-out area under the curve (AUC), which summarizes the tradeoff between sensitivity (fraction of correctly identified C19+ cases in the sample with obesity and without obesity) and specificity (fraction of correctly identified C19− cases in the sample with obesity and without obesity) as the threshold value for the predictor is varied. We used symptoms (binary), number of symptoms, chemosensory ratings during illness, COVID diagnosis, and days since onset of the respiratory illness. We focused on “during illness” ratings because those best showed evidence for the effects of illness and were also the most predictive symptom in a previous study with the same questionnaire (9). Moreover, this question (rather than pre-illness ratings or change in ratings) is best suited for being asked when making an inventory of symptoms in a clinical setting.

## RESULTS

### Participant Characteristics

A total of 5156 participants reported a positive lab test for COVID-19 (hereafter, C19+), while 659 reported a negative lab test for COVID-19 (hereafter, C19−). Of all participants, 519 (9% of the total group) self-reported to have obesity (C19+ = 433; C19− = 86) (**Figure 1**). The demographic profile of our participants is summarized in **Supplementary Table 2 a and b**. Age is higher in OB+ compared to OB− (43.1 vs 39.5) (BF_10_ = 7.48e+06 ± 0%). After excluding n = 17 participants with gender reporting categories of “prefer not to say” (n = 13) and “other” (n = 4), we observed different proportions of gender (χ^2^ = 4.42, p = 0.035), driven primarily by a higher proportion of women in the C19− group with obesity (87.2%) compared to those without obesity (77%).

### Before COVID-19 illness, participants with obesity exhibit similar smell, taste, and chemesthesis loss as those without obesity before COVID-19 illness

Before COVID-19 illness, OB+ participants did not self-report greater ability in smell (change against zero, BF_10_ = 6.48e-02 ± 0%), taste (BF_10_ = 7.24e-02 ± 0%) or chemesthesis (BF_10_ = 7.89e-02 ± 0%), or greater nasal congestion (BF_10_ = 6.80e-02 ± 0%) than OB− participants **(Supplementary Figure 1, Supplementary Table 3)**. Before COVID-19 illness C19+ participants reported greater ability in smell (BF_10_ = 2.16e+01) and taste (BF_10_ = 3.45e+02 ± 0%) than C19− participants.

### During COVID-19 illness, participants with obesity exhibit similar smell, taste, and chemesthesis loss as those without obesity during COVID-19 illness

C19+ participants reported greater deficits in smell (change against zero: BF_10_ = 1.20e+75 ± 0%), taste (change against zero: BF_10_ = 1.51e+29 ± 0%), and chemesthesis (change against zero: BF_10_ = 1.69e+05 ± 0%), as compared with C19− participants (**Figure 2, Supplementary Table 4**). Similar to our previous report (22), we reported lower deficits in nasal congestion with C19+ participants in our analysis (change against zero: BF_10_ = 1.56e+01 ± 0%), compared to C19− participants. Further, these chemosensory variables did not differ between the participants who self-reported obesity vs participants without obesity (smell BF_10_ = 1.44e-01 ± 0%; taste 5.7e-02 ± 0%; chemesthesis 8.22e-01 ± 0%), across the COVID groups. Similar to the above chemosensory findings during the illness, the differences in chemosensory ratings between pre-and during illness varied in C19+ and C19− participants (**Figure 3, Supplementary Table 5**). In particular, C19+ participants reported greater deficits in smell (change against zero: BF_10_ = 5.18e+59 ± 0%), taste (change against zero: BF_10_ =1.46e+32 ± 0%), and chemesthesis (change against zero: BF_10_ = 6.82e-+07 ± 0%), than the C19− group. However, when estimating the effect of obesity condition, the three chemosensory variables did not differ between the groups with obesity and without obesity across the COVID-19 condition (smell change against zero; BF_10_ = 6.71e-02 ± 0%; taste change against zero BF_10_ = 5.28e-02 ± 0%; chemesthesis change against zero BF_10_ = 1.26e-01 ± 0%). Interestingly, there was no main effect of COVID-19 condition or obesity status on the nasal obstruction reporting.

**Figure 2.**
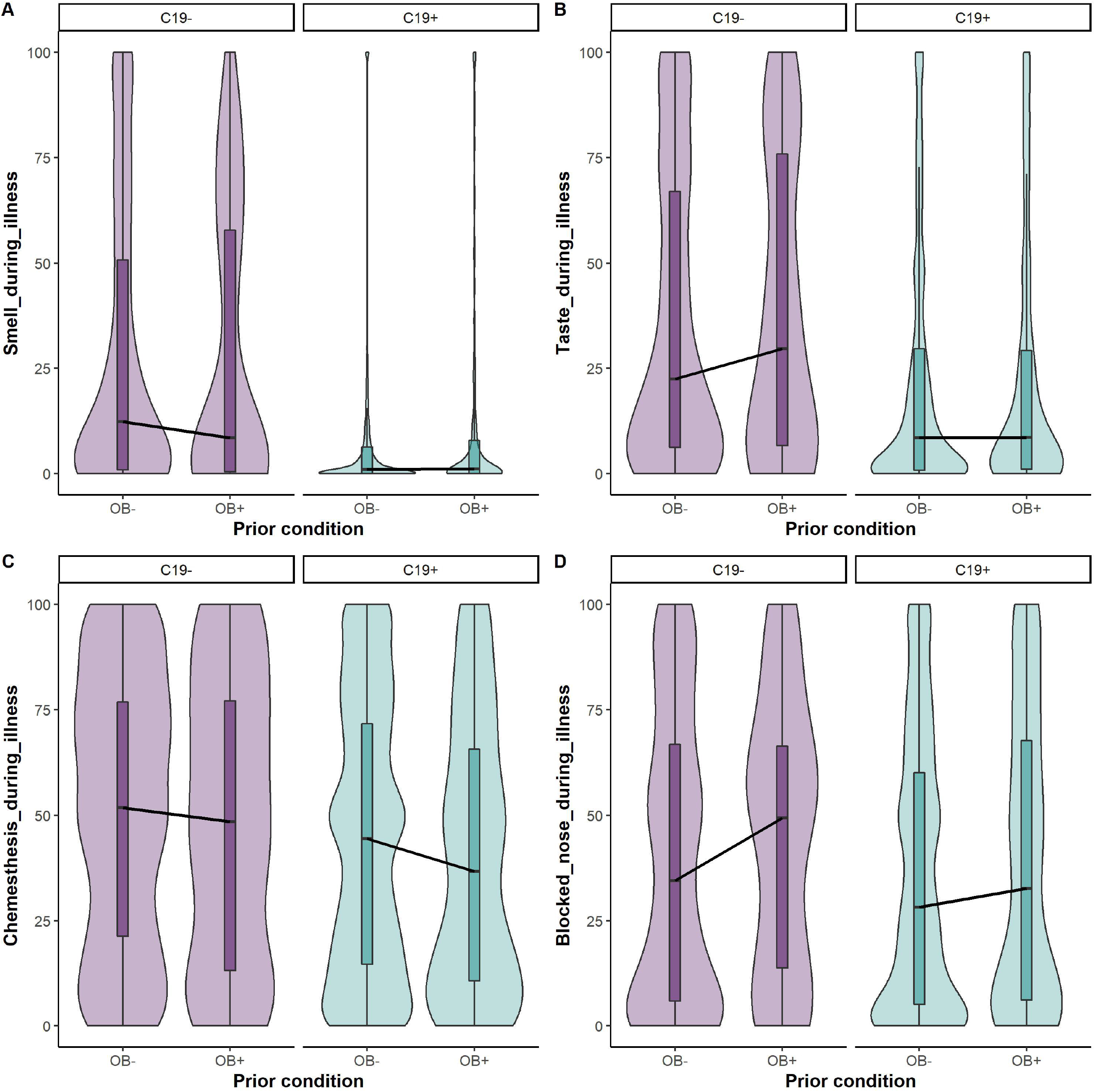
Self-reported smell (A), taste (B), chemesthesis (C), and nasal obstruction (D) ratings during the illness in C19+ (in purple) and C19− (in blue) participants with obesity (OB+) or without obesity (OB−). Ratings were given on 0-100 visual analog scales. Nasal obstruction question was formulated as “How blocked was your nose?”) during respiratory illness in C19+ and C19− participants. Each panel presents the mean ratings for chemosensory abilities and nasal blockage. All participants had a diagnosis via a lab test. The thick black horizontal bar indicates the median, the shaded bars within each violin indicates the interquartile range. The shaded violin area in purple and blue represents smoothed histogram of data density along the data points.

**Figure 3.**
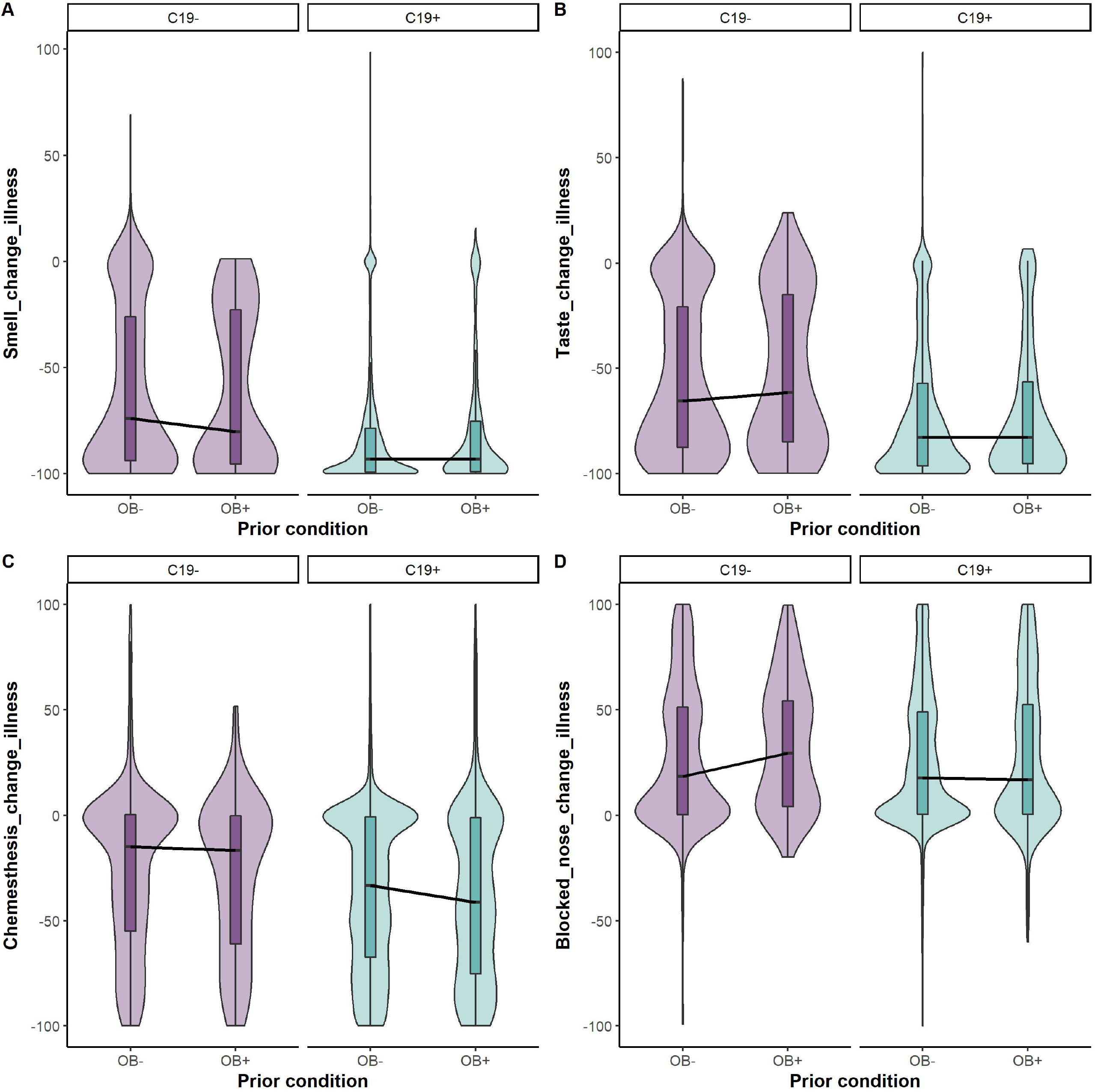
Self-reported change in smell (A), taste (B), chemesthesis (C), and nasal obstruction (D) ratings in C19+ (in purple) and C19− (in blue) participants with obesity (OB+) or without obesity (OB−). Each panel presents the distribution of the change scores, i.e., the rating “before” illness minus the rating “during” illness on the 100-point visual analog scale. All participants had a diagnosis via a lab test. The thick black horizontal bar indicates the median, the shaded bars within each violin indicates the interquartile range. The shaded violin area in purple and blue represents smoothed histogram of data density along the data points.

### Participants with obesity exhibit similar smell, taste, and chemesthesis recovery from COVID-19 illness as those without obesity

To further understand changes in chemosensory perception with COVID-19 diagnosis and obesity condition, we looked at the data from participants who reported recovery from the illness (**Figure 4, Supplementary Table 6**). Recovery was reported by 3970 participants (n=3431 C19+ and n= 539 C19−), which is approximately 68% of our sample. Our Bayesian linear models suggest that the ratings for post-recovery chemosensory perception (smell BF_10_ = 7.79e-02 ± 0.02%; taste BF_10_ = 6.44e-02 ± 0.02%; chemesthesis BF_10_ = 2.18e-01 ± 0.01%) did not differ in C19+ and C19− diagnosis. Of note, some smell/taste/chemosensory symptoms remain post-recovery from the illness in C19+ and C19− groups. We found no differences in smell (BF_10_ = 6.58e-02 ± 0.02%), taste (BF_10_ = 1.07e-01 ± 0.02%), and chemesthetic perception (BF_10_ = 9.96e-02 ± 0.11%) by self-reported obesity. Nasal obstruction did not seem to be affected by either COVID-19 diagnosis (BF_10_ = 5.33e-02 ± 0.03%) or obesity status (BF_10_ =8.16e-02 ± 0.02%) post-recovery from the illness.

**Figure 4.**
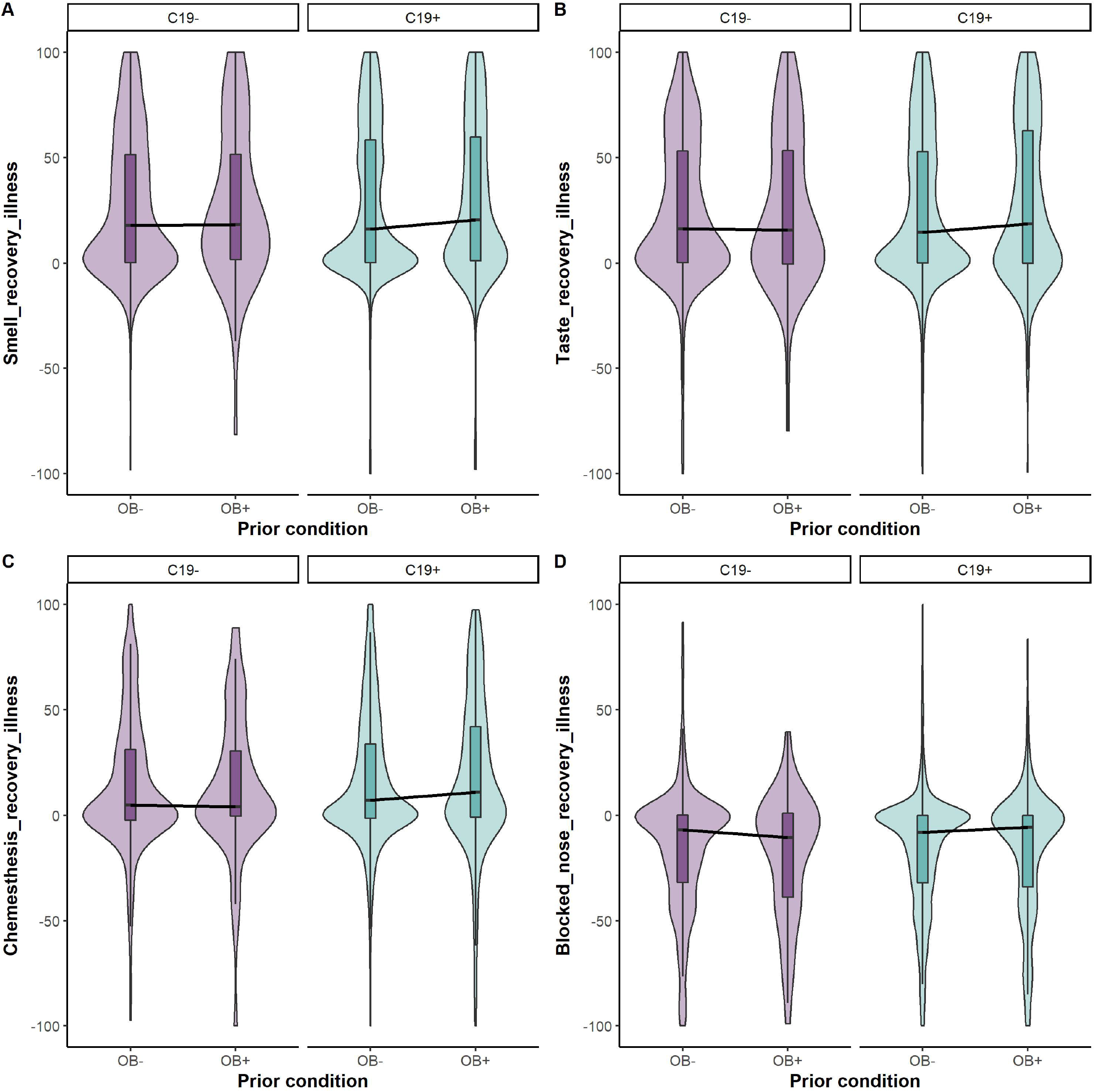
Self-reported change in smell (A), taste (B), chemesthesis (C), and nasal obstruction (D) ratings post-recovery from respiratory illness in C19+ (in purple) and C19− (in blue) participants with obesity (OB+) or without obesity (OB−). Ratings were given on 0-100 visual analog scales. Each panel presents the mean ratings for chemosensory abilities and nasal blockage post-recovery from respiratory illness. All participants had a diagnosis via a lab test. The thick black horizontal bar indicates the median, the shaded bars within each violin indicates the interquartile range. The shaded violin area in purple and blue represents smoothed histogram of data density along the data points.

### Participants with obesity report more symptoms overall and more frequently report respiratory and gastrointestinal (GI) symptoms

Based on the evidence from existing clinical and epidemiological studies, one of our goals was to assess whether individuals with self-reported obesity overall have greater symptomatic manifestation with C19+ diagnosis than those participants without obesity. To test our hypothesis, we used Bayesian linear regression and compared the sum of the symptoms reported by participants in these samples versus samples without obesity **(Figure 5A, Supplementary Table 8**). As predicted, among those with C19+, there is decisive evidence that participants with obesity report a larger number of symptoms than participants without obesity (BF_10_ = 1.02e04 ±0%; average N of symptoms = with obesity: 8.22; without obesity: 7.42). A similar effect is observed among participants with C19− (average N of symptoms = with obesity: 8; without obesity: 7.33 BF_10_ = 9.91e03 ±0%). Among those with C19+, disease duration is longer in those with obesity (BF_10_ = 1.02e04 ±0%; average days since onset), while in C19− such a difference is not observed (BF_10_ = 1.21e00 ±0% (**Figure 5B, Supplementary Table 7**). Looking at the specific symptoms (**Figure 5C**), smell and taste symptoms are equally reported by participants with obesity and participants without obesity with a diagnosis of COVID-19. Further, participants with self-reported obesity reported greater frequency in loss of appetite, diarrhea, and nausea, along with shortness of breath, cough (dry or with mucus), and chest tightness.

**Figure 5.**
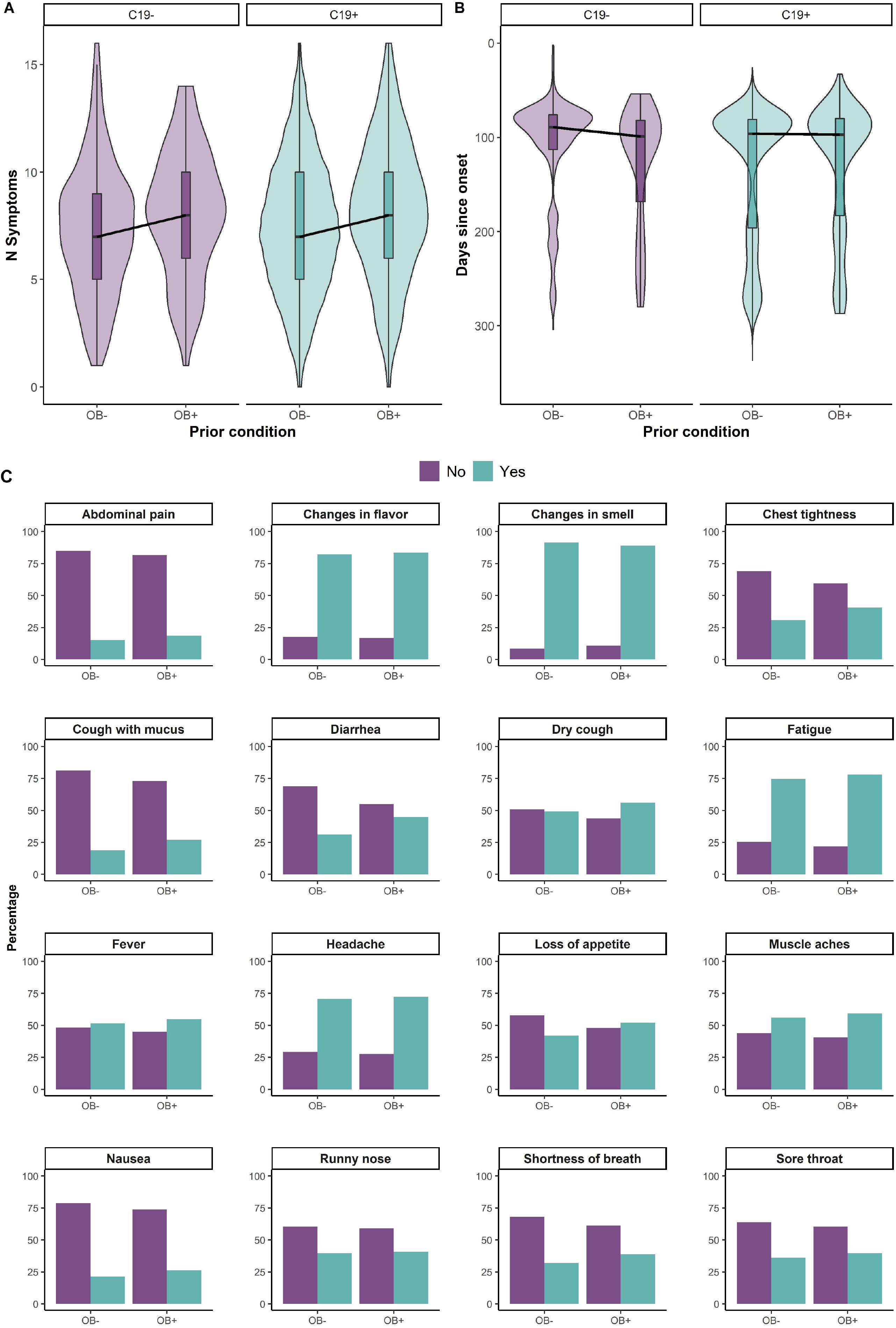
Self-reported symptomatic manifestation reported by C19+ (in purple) and C19− (in blue) participants with obesity (OB+) or without obesity (OB−). **(A)** Cumulative number of symptoms reported by C19+ (in purple) and C19− (in blue) participants with obesity (OB+) or without obesity (OB−). **(B)** Self-reported average number of days since onset of respiratory illness symptoms reported by C19+ (in purple) and C19− (in blue) participants with obesity (OB+) or without obesity (OB−). **(C)** Proportion of participants with C19+ that report specific symptoms by self-reported obesity (OB+) or without obesity (OB−). * p<0.05. The thick black horizontal bar indicates the median, the shaded bars within each violin indicates the interquartile range. The shaded violin area in purple and blue represents smoothed histogram of data density along the data points.

### A classifier trained on participants without obesity accurately predicts C19+ diagnosis in participants with obesity

Based on the self-reports on symptoms, combined with the chemosensory and nasal obstruction ratings, we assessed the accuracy with which we could predict a C19+ diagnosis (**Figure 6**) in OB−. We then tested the model to predict the accuracy of discrimination of C19+ in participants with obesity. Our results indicate that we can predict the C19+ diagnosis with 63% accuracy, which indicates a moderately good estimate. Variables included in this analysis are reported in **Supplementary Table 8.**

**Figure 6.**
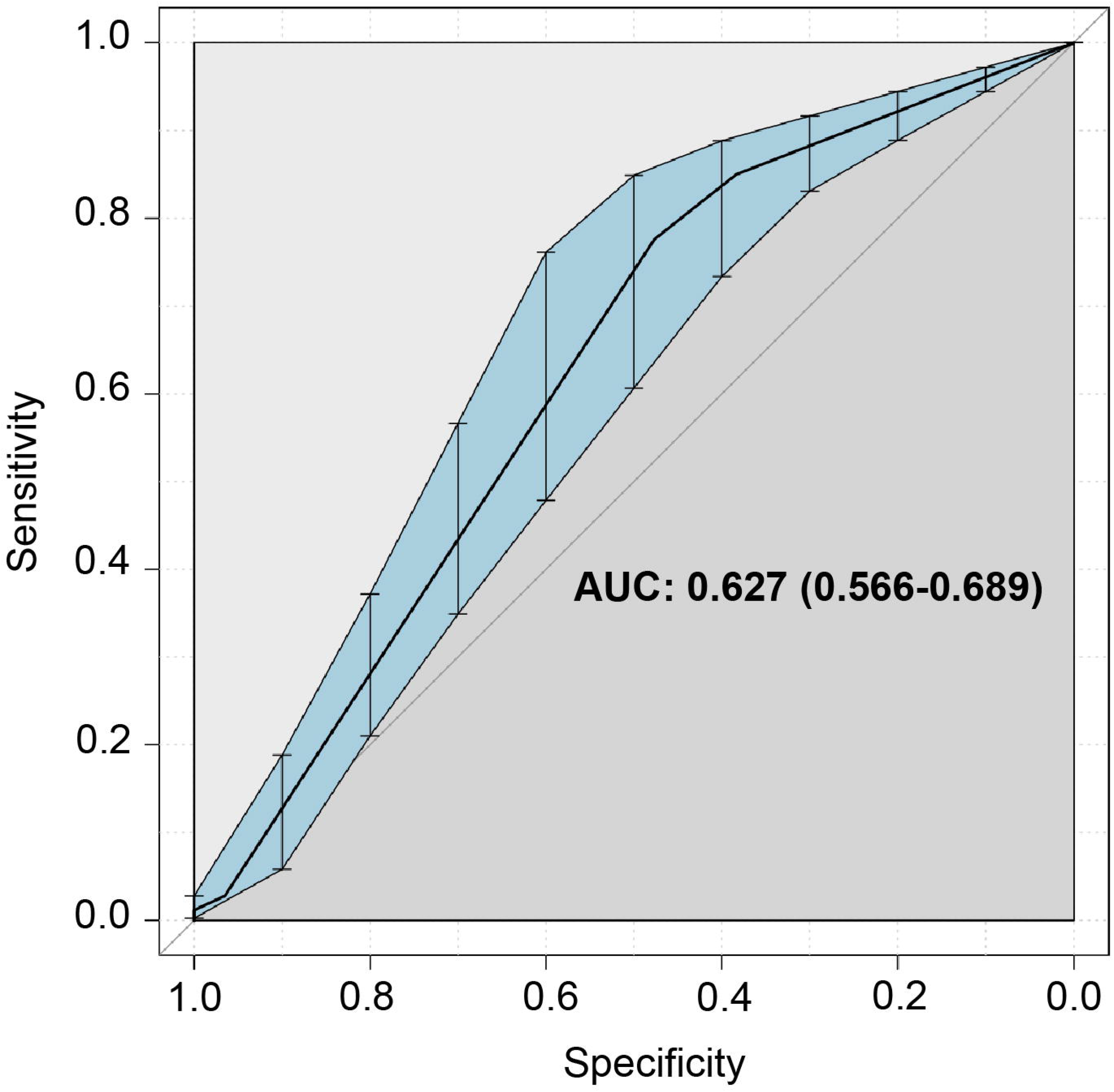
ROC curve in discriminating C19+ vs. C19− in participants with obesity (OB+) after having trained the model with participants without obesity (OB−).

## DISCUSSION

Since the beginning of the COVID-19 pandemic, reports of olfactory and gustatory dysfunctions in COVID-19 patients continue to grow. To our knowledge, this study is the first to describe and compare the chemosensory perception and related symptomatology in COVID-19 patients who self-reported to have obesity vs. no obesity. Independent of the obesity status, the subjective ratings of smell, taste, and chemesthesis declined with COVID-19 illness. Examining the recovery patterns, we found that participants with obesity show similar recovery from COVID-19 related loss of smell, taste, and chemesthesis as those without obesity. Although we do not know the severity of each symptom, those with obesity reported a greater frequency of respiratory and GI symptoms and more symptoms overall. Finally, we found that a model of all symptoms combined that was trained on patients without obesity is, can predict the C19+ diagnosis with 63% of accuracy in participants with obesity. Furthermore, this smell loss was not related to self-reported nasal obstruction, commonly observed in other upper respiratory infections (33,34). Together, these results confirm and add to previous reports that COVID-19 largely impacts chemosensory function; however, obesity does not mask self-reported chemosensory loss in those with a COVID diagnosis.

Smell and taste disturbances are a typical consequence of nasal inflammation due to an upper respiratory tract viral infection (35,36); however, an acute loss of taste and smell emerged rapidly as a critical neurological manifestation of a positive COVID-19 diagnosis (37). Our current findings are similar to prior reports that showed that approximately 90% of the participants reported a loss of smell. Furthermore, nearly 80% of the participants reported a loss of taste, and 46% had a reduction of chemesthesis (detection of chemicals that induce tingling and burning sensations such as the burning of chili peppers), indicating that the chemosensory impairment is not restricted to smell (9,22). While most cold viruses cause nasal congestion and individuals experience a reduction in the sense of smell, our results showed that nasal congestion was not associated with smell loss. This finding is consistent with other reports (38–40) where individuals with COVID-19 do not report clinically significant nasal congestion or rhinorrhea, suggesting that other mechanisms may play a role in COVID-19 associated smell loss (37).

In addition to being a risk factor for COVID-19 viral infection, excessive body weight is also implicated in chemosensory decline. Adipose tissue in obesity is “pro-inflammatory”, causing a surge in levels of IL-6 and C-reactive protein and enhancing the expression of cytokines and adipokines (41). Interestingly, in diseases where these circulating inflammatory factors are high, smell and taste dysfunction are prevalent (42,43). In particular, acute induction of systemic inflammation has been shown to shorten the lifespan of adult taste bud cells (18).

Similarly, enhanced expression of inflammatory markers is shown to reduce olfactory sensory neurons, in mice fed a high-fat diet to induce obesity (44). Thus, obesity-related inflammation may affect chemosensory function. A major concern with this pre-existing gustatory and olfactory sensory deficiency due to obesity is that obesity may mask the viral-induced diminished taste and smell self-reported experiences. Interestingly, our analysis showed that COVID-19 related chemosensory-related changes were comparable between C19+ participants with obesity and without obesity suggesting that obesity does not have an effect on the loss of chemosensory perception with COVID diagnosis. These findings need to be taken with caution, especially when considering severe cases, which are more common in patients with obesity. For example, if a patient is in critical condition, they cannot pay attention to their chemosensory alterations, and chemosensory perception will likely not be tested or self-reported. This does not mean that the chemosensory perception is not affected.

In terms of chemosensory recovery, we found no differences between participants with obesity compared to those without obesity. While none of the studies to date have compared the recovery rates between C19+ participants with obesity vs no obesity, our overall recovery rate of 65% is comparable to our previous analysis (9) but slightly lower than other studies (45,46). There are residuals smell/taste/chemosensory symptoms reported post-recovery from the illness in C19+ and C19− groups. In particular, quantitative studies using psychophysical methods have shown that nearly 25% of people continue to report chemosensory problems when evaluated 30 - 60 days after the onset of COVID-19 (45). This insufficient recovery rate may significantly increase the number of patients with chemosensory disturbances, ultimately influencing eating behaviors (47), quality of life (48,49), and psychological health (50,51) in the general population. But most importantly, it may significantly impact participants with obesity who have an added burden of lower chemosensory acuity due to excess fat mass (44,52). Thus, it is imperative to prepare healthcare workers to detect and treat chemosensory disorders in this high-risk population.

As we hypothesized, non-chemosensory symptoms were more severe in C19+ participants with obesity than in participants without obesity. Specifically, participants with obesity reported a greater frequency of respiratory and GI symptoms. In general, it is known that obesity is associated with GI symptoms disturbances, such as upper abdominal pain, nausea, vomiting, retching, and gastritis. GI symptoms are accompanied by inflammation or alterations of intestinal permeability (53–56). However, it also emerged that COVID-19 patients experienced several GI symptoms such as diarrhea (24.2%), anorexia (17.9%), and nausea (17.9%) (57), though they vary widely and are less understood. However, this may not be surprising since some viral infections are known to cause alterations in intestinal permeability as well (58). The mediation of ACE2 cell receptors could elucidate the mechanism related to GI tract involvement in SARS-CoV-2 infection. While ACE2 is expressed in abundance in the lungs’ alveolar cells, the receptor is also highly expressed in the GI tract, especially in the small and large intestines (59–63).

Our study has some limitations. Our online survey and sampling methodology likely selected participants with a heightened interest in smell and taste and/or their disturbances. Due to that, the data collected at the peak of the pandemic obesity was self-reported; thus, we acknowledge the potential under-reporting of obesity. We also acknowledge that due to the nature of our data being collected in several countries, the definition of obesity may vary and there may be regional and cultural factors that may influence stigma and biases towards self-report of obesity. Ideally, future studies using quantitative taste and smell measures will be conducted in this population. However, although the taste and smell reports were also self-reports, similar to prior studies, we demonstrate that self-reported taste and smell may be a helpful tool to distinguish between C19+ and C19−.

Despite the limitations, our study shows differences in participants with obesity compared to participants without obesity with other symptoms. However, those differences potentially do not affect the chemosensory symptoms. In general, more evidence is needed to understand biological mechanisms related to alterations in taste and smell loss in individuals with COVID-19. Understanding how the alteration initiates and progresses will provide molecular and cellular bases for diagnosis and treatment of chemosensory disorders for those with COVID-19 and others who lose their sense of taste and smell due to other conditions with underlying inflammation. While it is an exciting prospect, the use of chemosensory assessments as an effective tool for screening and treatment protocols, and the possibility of integrating these tests into current COVID-19 screening protocols have yet to be determined in the general population, as well as high-risk populations with obesity.

## Data Availability

The research compendium including data files and analysis scripts is available at data in OSF

https://osf.io/rbcty

## Acknowledgments

The authors wish to thank all study participants, patients, and patient advocates that have contributed to this project.

