## Supplementary material for "COVID-19 Related Chemosensory Changes in Individuals with Self-Reported Obesity": Supplentary Material

**Supplementary Information**

**Supplementary Table 1.** Interpretation of Bayes factors BF_10_ following the classification proposed by Lee and Wagenmakers (2013) and adjusted from Jeffreys (1961)

| **Bayes factor** | **Evidence category** |
| --- | --- |
| >100 | Extreme evidence for H_1_ |
| 30–100 | Very strong evidence for H_1_ |
| 10–30 | Strong evidence for H_1_ |
| 3–10 | Moderate evidence for H_1_ |
| 1–3 | Anecdotal evidence for H_1_ |
| 1 | No evidence |
| 1/3–1 | Anecdotal evidence for H_0_ |
| 1/10–1/3 | Moderate evidence for H_0_ |
| 1/30–1/10 | Strong evidence for H_0_ |
| 1/100–1/30 | Very strong evidence for H_0_ |
| <1/100 | Extreme evidence for H_0_ |

**Supplementary Table 2a.** Participant gender between C19+ and C19- participants who self-reported to have obesity (OB+) or no obesity (OB-).

|  | **All** | | **C19+** | | **C19-** | |
| --- | --- | --- | --- | --- | --- | --- |
|  | **OB+** | **OB-** | **OB+** | **OB-** | **OB+** | **OB-** |
| Women, n | 412 | 3571 | 337 | 3165 | 75 | 406 |
| Women proportion | 0.793834 | 0.750999 | 0.778291 | 0.748581 | 0.872093 | 0.770398 |
| Men, n | 107 | 1184 | 96 | 1063 | 11 | 121 |
| Men proportion | 0.206166 | 0.249001 | 0.221709 | 0.251419 | 0.127907 | 0.229602 |
| Chi Sq statistic (p-value) | 4.4155 (0.03561) | | 1.7001 (0.1923) | | 3.9434 (0.04705) | |

C19+ = positive COVID-19 diagnosis; C19- = negative COVID-19 diagnosis; OB+ = with obesity; OB+ = without obesity

**Supplementary Table 2b.** Participant age between C19+ and C19- participants who self-reported to have obesity (OB+) or no obesity (OB-).

| **Model (lmBF, on data and with random effect of ID)** | **Bayes Factor (JZS)** | **± error%** |
| --- | --- | --- |
| Age ~ Group+Obesity+Group:Obesity | 2.60E+10 | 1.46E-02 |
| Age ~ Group+Obesity | 5.63E+10 | 1.02E-02 |
| Age ~ Group | 7.52E+03 | 1.62E-12 |
| Age ~ Obesity | 3.07E+07 | 1.48E-16 |
| Effect of Obesity:(X ~ Group+Obesity)/ (X ~ Group) | 7.48E+06 | 1.02E-02 |
| Effect of Group:(X ~ Group+Obesity)/(X ~ Obesity) | 1.83E+03 | 1.02E-02 |

C19+ = positive COVID-19 diagnosis; C19- = negative COVID-19 diagnosis; OB+ = with obesity; OB+ = without obesity. lmBF = Function to compute Bayes Factors for specific linear models. JZS = JeffreysZellner-Siow priors

**Supplementary Table 3.** Chemosensory and nasal obstruction ratings before the respiratory illness in C19+ and C19- participants who self-reported to have obesity (OB+) or no obesity (OB-).

|  | **Model (lmBF, on data and with random effect of ID)** | **Bayes Factor (JZS)** | **± error%** |
| --- | --- | --- | --- |
| Smell | Smell_before_illness ~ Group+Obesity+Age+Group:Obesity | 1.46E+00 | 2.84E-01 |
|  | Smell_before_illness ~ Group+Obesity+Age | 8.94E+00 | 1.59E-02 |
|  | Smell_before_illness ~ Group+Age | **1.38E+02** | 3.37E-02 |
|  | Smell_before_illness ~ Obesity+Age | 4.14E-01 | 2.17E-02 |
|  | **Effect of Obesity:(X ~ Group+Obesity+Age)/ (X ~ Group+Age)** | 6.48E-02 | 3.72E-02 |
|  | Effect of Group:(X ~ Group+Obesity+Age)/(X ~ Obesity+Age) | **2.16E+01** | 2.69E-02 |
| Taste | Taste_before_illness ~ Group+Obesity+Age+Group:Obesity | 9.37E+00 | 2.09E-02 |
|  | Taste_before_illness ~ Group+Obesity+Age | **8.88E+01** | 2.42E-02 |
|  | Taste_before_illness ~ Group+Age | **1.23E+03** | 2.29E-02 |
|  | Taste_before_illness ~ Obesity+Age | 2.57E-01 | 1.18E-02 |
|  | **Effect of Obesity:(X ~ Group+Obesity+Age)/ (X ~ Group+Age)** | 7.24E-02 | 3.33E-02 |
|  | Effect of Group:(X ~ Group+Obesity+Age)/(X ~ Obesity+Age) | **3.45E+02** | 2.69E-02 |
| Chemesthesis | Chemesthesis_before_illness ~ Group+Obesity+Age+Group:Obesity | 7.86E-04 | 2.49E-02 |
|  | Chemesthesis_before_illness ~ Group+Obesity+Age | 6.99E-03 | 2.07E-02 |
|  | Chemesthesis_before_illness ~ Group+Age | 8.86E-02 | 1.18E-02 |
|  | Chemesthesis_before_illness ~ Obesity+Age | 4.89E-03 | 1.45E-02 |
|  | **Effect of Obesity:(X ~ Group+Obesity+Age)/ (X ~ Group+Age)** | 7.89E-02 | 2.38E-02 |
|  | Effect of Group:(X ~ Group+Obesity+Age)/(X ~ Obesity+Age) | 1.43E+00 | 2.52E-02 |
| Blocked nose | Blocked_nose_before_illness ~ Group+Obesity+Age+Group:Obesity | 6.31E-04 | 2.20E-02 |
|  | Blocked_nose_before_illness ~ Group+Obesity+Age | 6.45E-03 | 3.29E-02 |
|  | Blocked_nose_before_illness ~ Group+Age | 9.48E-02 | 1.17E-02 |
|  | Blocked_nose_before_illness ~ Obesity+Age | 7.64E-03 | 1.87E-02 |
|  | **Effect of Obesity:(X ~ Group+Obesity+Age)/ (X ~ Group+Age)** | 6.80E-02 | 3.49E-02 |
|  | Effect of Group:(X ~ Group+Obesity+Age)/(X ~ Obesity+Age) | 8.44E-01 | 3.79E-02 |

Ratings were given on 0-100 visual analog scales. In the second column “model”, the effect of Obesity is made bold, because that is our most important model. In column “BF” any value for strong evidence (BF>10) for H_1_ is bold, any value indicative of strong evidence for H0 (no difference) is underlined. lmBF = Function to compute Bayes Factors for specific linear models. JZS = JeffreysZellner-Siow priors

**Supplementary Figure 1.** Self-reported smell (A), taste (B), chemesthesis (C), and nasal obstruction (D) ratings before the respiratory illness in C19+ (in purple) and C19- (in blue) participants with obesity (OB+) or without obesity (OB-). Ratings were given on 0-100 visual analog scales. Nasal obstruction question was formulated as “How blocked was your nose?”) before the respiratory illness in C19+ and C19- participants. Each panel presents the mean ratings for chemosensory abilities and nasal blockage. All participants had a diagnosis via a lab test. The thick black horizontal bar connects medians, the shaded bars within each violin indicates the interquartile range. The shaded violin area in purple and blue represents smoothed histogram of data density along the data points.

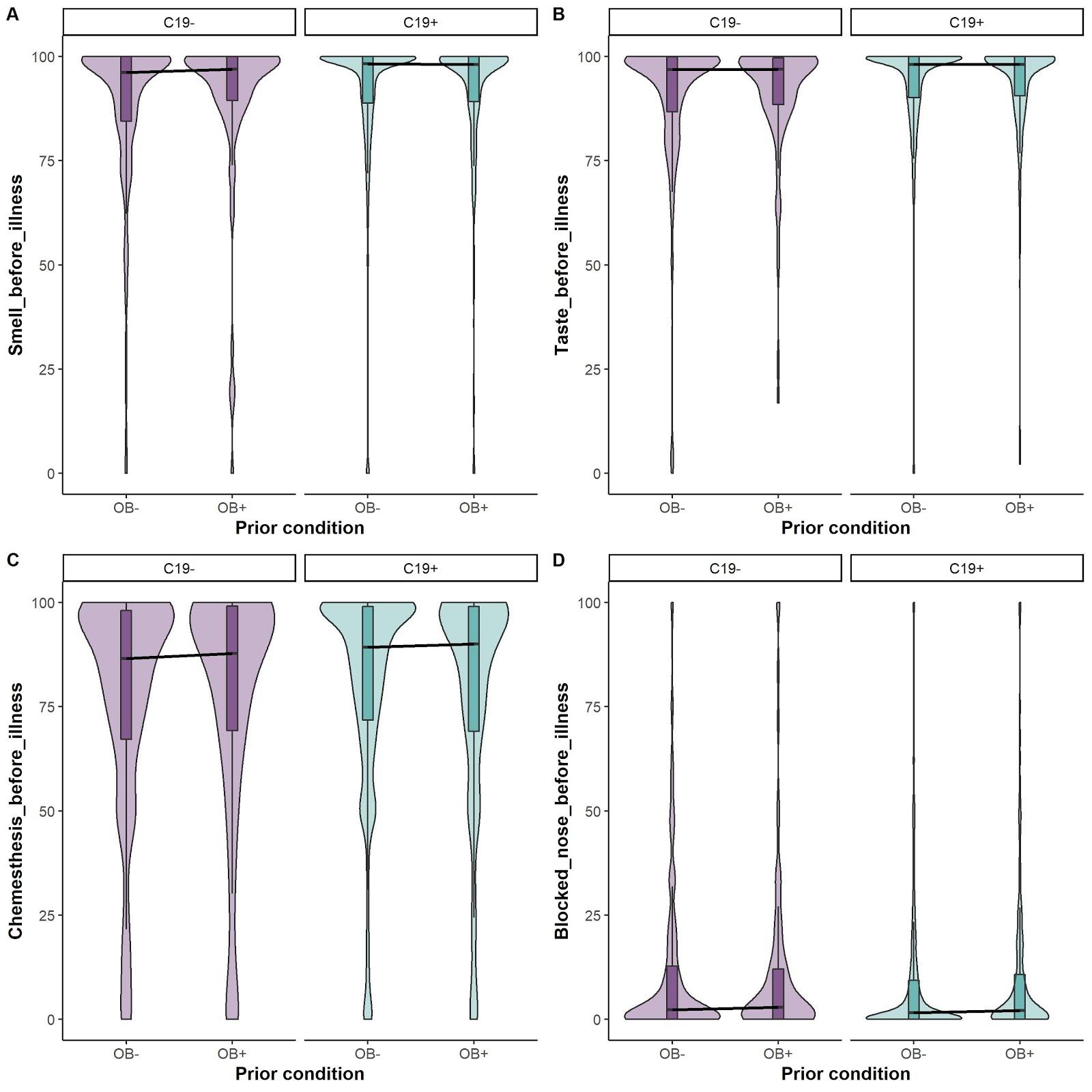

**Supplementary Table 4.** Chemosensory and nasal obstruction ratings during the respiratory illness in C19+ and C19- participants who self-reported to have obesity (OB+) or no obesity (OB-).

|  | **Model (lmBF, on data and with random effect of ID)** | **Bayes Factor (JZS** | **± error%** |
| --- | --- | --- | --- |
| Smell | Smell_during_illness ~ Group+Obesity+Age+Group:Obesity | **1.76E+84** | 3.54E-02 |
|  | Smell_during_illness ~ Group+Obesity+Age | **1.33E+85** | 1.36E-02 |
|  | Smell_during_illness ~ Group+Age | **9.23E+85** | 1.71E-02 |
|  | Smell_during_illness ~ Obesity+Age | **1.11E+10** | 1.53E-02 |
|  | **Effect of Obesity:(X ~ Group+Obesity+Age)/ (X ~ Group+Age)** | 1.44E-01 | 2.19E-02 |
|  | Effect of Group:(X ~ Group+Obesity+Age)/(X ~ Obesity+Age) | **1.20E+75** | 2.05E-02 |
| Taste | Taste_during_illness ~ Group+Obesity+Age+Group:Obesity | **5.20E+25** | 5.23E-02 |
|  | Taste_during_illness ~ Group+Obesity+Age | **4.31E+26** | 2.39E-02 |
|  | Taste_during_illness ~ Group+Age | **7.57E+27** | 9.70E-03 |
|  | Taste_during_illness ~ Obesity+Age | 2.85E-03 | 1.58E-02 |
|  | **Effect of Obesity:(X ~ Group+Obesity+Age)/ (X ~ Group+Age)** | 5.70E-02 | 2.58E-02 |
|  | Effect of Group:(X ~ Group+Obesity+Age)/(X ~ Obesity+Age) | **1.51E+29** | 2.87E-02 |
| Chemesthesis | Chemesthesis_during_illness ~ Group+Obesity+Age+Group:Obesity | **1.83E+09** | 3.83E-02 |
|  | Chemesthesis_during_illness ~ Group+Obesity+Age | **1.85E+10** | 1.52E-02 |
|  | Chemesthesis_during_illness ~ Group+Age | **2.25E+10** | 1.27E-02 |
|  | Chemesthesis_during_illness ~ Obesity+Age | **1.09E+05** | 4.24E-02 |
|  | **Effect of Obesity:(X ~ Group+Obesity+Age)/ (X ~ Group+Age)** | 8.22E-01 | 1.98E-02 |
|  | Effect of Group:(X ~ Group+Obesity+Age)/(X ~ Obesity+Age) | **1.69E+05** | 4.50E-02 |
| Blocked nose | Blocked_nose_during_illness ~ Group+Obesity+Age+Group:Obesity | **2.31E+07** | 5.67E-02 |
|  | Blocked_nose_during_illness ~ Group+Obesity+Age | **2.03E+08** | 1.52E-02 |
|  | Blocked_nose_during_illness ~ Group+Age | **5.97E+07** | 3.48E-02 |
|  | Blocked_nose_during_illness ~ Obesity+Age | **1.30E+07** | 1.11E-02 |
|  | **Effect of Obesity:(X ~ Group+Obesity+Age)/ (X ~ Group+Age)** | 3.40E+00 | 3.80E-02 |
|  | Effect of Group:(X ~ Group+Obesity+Age)/(X ~ Obesity+Age) | **1.56E+01** | 1.88E-02 |

Ratings were given on 0-100 visual analog scales. In the second column “model”, the effect of Obesity is made bold, because that is our most important model. In column “BF” any value for strong evidence (BF>10) for H_1_ is bold, any value indicative of strong evidence for H0 (no difference) is underlined. lmBF = Function to compute Bayes Factors for specific linear models. JZS = JeffreysZellner-Siow priors

**Supplementary Table 5.** Change in chemosensory and nasal obstruction ratings in C19+ (in purple) and C19- (in blue) participants who self-reported to have obesity (OB+) or no obesity (OB-).

|  | **Model (lmBF, on data and with random effect of ID)** | **Bayes Factor (JZS)** | **± error%** |
| --- | --- | --- | --- |
| Smell | Smell_change_illness ~ Group+Obesity+Age+Group:Obesity | **3.75E+69** | 3.61E-02 |
|  | Smell_change_illness ~ Group+Obesity+Age | **2.51E+70** | 1.33E-02 |
|  | Smell_change_illness ~ Group+Age | **3.74E+71** | 1.86E-02 |
|  | Smell_change_illness ~ Obesity+Age | **4.85E+10** | 1.44E-01 |
|  | **Effect of Obesity:(X ~ Group+Obesity+Age)/ (X ~ Group+Age)** | 6.71E-02 | 2.29E-02 |
|  | Effect of Group:(X ~ Group+Obesity+Age)/(X ~ Obesity+Age) | **5.18E+59** | 1.45E-01 |
| Taste | Taste_change_illness ~ Group+Obesity+Age+Group:Obesity | **1.39E+29** | 3.20E-01 |
|  | Taste_change_illness ~ Group+Obesity+Age | **9.56E+29** | 1.46E-02 |
|  | Taste_change_illness ~ Group+Age | **1.81E+31** | 9.62E-03 |
|  | Taste_change_illness ~ Obesity+Age | 6.55E-03 | 3.47E-02 |
|  | **Effect of Obesity:(X ~ Group+Obesity+Age)/ (X ~ Group+Age)** | 5.28E-02 | 1.75E-02 |
|  | Effect of Group:(X ~ Group+Obesity+Age)/(X ~ Obesity+Age) | **1.46E+32** | 3.77E-02 |
| Chemesthesis | Chemesthesis_change_illness ~ Group+Obesity+Age+Group:Obesity | **8.90E+07** | 2.96E-02 |
|  | Chemesthesis_change_illness ~ Group+Obesity+Age | **9.45E+08** | 1.51E-02 |
|  | Chemesthesis_change_illness ~ Group+Age | **7.48E+09** | 2.93E-02 |
|  | Chemesthesis_change_illness ~ Obesity+Age | **1.39E+01** | 1.68E-02 |
|  | **Effect of Obesity:(X ~ Group+Obesity+Age)/ (X ~ Group+Age)** | 1.26E-01 | 3.29E-02 |
|  | Effect of Group:(X ~ Group+Obesity+Age)/(X ~ Obesity+Age) | **6.82E+07** | 2.26E-02 |
| Blocked nose | Blocked_nose_change_illness ~ Group+Obesity+Age+Group:Obesity | **1.97E+07** | 2.58E-02 |
|  | Blocked_nose_change_illness ~ Group+Obesity+Age | **1.76E+08** | 3.70E-02 |
|  | Blocked_nose_change_illness ~ Group+Age | **1.88E+08** | 2.89E-02 |
|  | Blocked_nose_change_illness ~ Obesity+Age | **4.64E+08** | 1.13E-02 |
|  | **Effect of Obesity:(X ~ Group+Obesity+Age)/ (X ~ Group+Age)** | 9.34E-01 | 4.70E-02 |
|  | Effect of Group:(X ~ Group+Obesity+Age)/(X ~ Obesity+Age) | 3.79E-01 | 3.87E-02 |

Ratings were given on 0-100 visual analog scales. In the second column “model”, the effect of Obesity is made bold, because that is our most important model. In column “BF” any value for strong evidence (BF>10) for H_1_ is bold, any value indicative of strong evidence for H0 (no difference) is underlined. lmBF = Function to compute Bayes Factors for specific linear models. JZS = JeffreysZellner-Siow priors

**Supplementary Table 6.** Chemosensory and nasal obstruction ratings post-recovery from respiratory illness in C19+ and C19- participants who self-reported to have obesity (OB+) or no obesity (OB-).

|  | **Model (lmBF, on data and with random effect of ID)** | **Bayes Factor (JZS)** | **± error%** |
| --- | --- | --- | --- |
| Smell | Smell_recovery_illness ~ Group+Obesity+Age+Group:Obesity | 5.85E-05 | 4.32E-02 |
|  | Smell_recovery_illness ~ Group+Obesity+Age | 4.81E-04 | 1.71E-02 |
|  | Smell_recovery_illness ~ Group+Age | 7.31E-03 | 1.49E-02 |
|  | Smell_recovery_illness ~ Obesity+Age | 6.18E-03 | 1.22E-02 |
|  | **Effect of Obesity:(X ~ Group+Obesity+Age)/ (X ~ Group+Age)** | 6.58E-02 | 2.27E-02 |
|  | Effect of Group:(X ~ Group+Obesity+Age)/(X ~ Obesity+Age) | 7.79E-02 | 2.10E-02 |
| Taste | Taste_recovery_illness ~ Group+Obesity+Age+Group:Obesity | 5.84E-05 | 1.13E-01 |
|  | Taste_recovery_illness ~ Group+Obesity+Age | 3.72E-04 | 2.43E-02 |
|  | Taste_recovery_illness ~ Group+Age | 3.47E-03 | 1.55E-02 |
|  | Taste_recovery_illness ~ Obesity+Age | 5.77E-03 | 4.08E-02 |
|  | **Effect of Obesity:(X ~ Group+Obesity+Age)/ (X ~ Group+Age)** | 1.07E-01 | 2.88E-02 |
|  | Effect of Group:(X ~ Group+Obesity+Age)/(X ~ Obesity+Age) | 6.44E-02 | 4.75E-02 |
| Chemesthesis | Chemesthesis_recovery_illness ~ Group+Obesity+Age+Group:Obesity | 2.70E-04 | 3.63E-02 |
|  | Chemesthesis_recovery_illness ~ Group+Obesity+Age | 1.81E-03 | 8.19E-02 |
|  | Chemesthesis_recovery_illness ~ Group+Age | 1.82E-02 | 1.17E-02 |
|  | Chemesthesis_recovery_illness ~ Obesity+Age | 8.32E-03 | 1.18E-02 |
|  | **Effect of Obesity:(X ~ Group+Obesity+Age)/ (X ~ Group+Age)** | 9.96E-02 | 8.27E-02 |
|  | Effect of Group:(X ~ Group+Obesity+Age)/(X ~ Obesity+Age) | 2.18E-01 | 8.27E-02 |
| Blocked nose | Blocked_nose_recovery_illness ~ Group+Obesity+Age+Group:Obesity | 6.00E-01 | 2.19E-02 |
|  | Blocked_nose_recovery_illness ~ Group+Obesity+Age | 4.57E+00 | 2.14E-02 |
|  | Blocked_nose_recovery_illness ~ Group+Age | **5.59E+01** | 1.16E-02 |
|  | Blocked_nose_recovery_illness ~ Obesity+Age | **8.56E+01** | 2.17E-02 |
|  | **Effect of Obesity:(X ~ Group+Obesity+Age)/ (X ~ Group+Age)** | 8.16E-02 | 2.44E-02 |
|  | Effect of Group:(X ~ Group+Obesity+Age)/(X ~ Obesity+Age) | 5.33E-02 | 3.05E-02 |

Ratings were given on 0-100 visual analog scales. In the second column “model”, the effect of Obesity is made bold, because that is our most important model. In column “BF” any value for strong evidence (BF>10) for H_1_ is bold, any value indicative of strong evidence for H0 (no difference) is underlined. lmBF = Function to compute Bayes Factors for specific linear models. JZS = JeffreysZellner-Siow priors

**Supplementary Table 7.** Severity of symptoms and days since onset in C19+ (in purple) and C19- (in blue) participants with obesity (OB+) or without obesity (OB-).

| **Model (lmBF, on data and with random effect of ID)** | **Bayes Factor (JZS)** | **± error%** |
| --- | --- | --- |
| N symptoms ~ Obesity+Age | **3.01E+03** | 1.11E-02 |
| N symptom ~ Obesity | 2.95E-01 | 1.84E-05 |
| Effect of Obesity:(X ~ Obesity+Age)/ (X ~ Age) | **1.02E+04** | 1.11E-02 |
| N symptoms ~ Obesity+Age | **2.92E+03** | 1.11E-02 |
| N symptom ~ Obesity | 2.95E-01 | 1.84E-05 |
| Effect of Obesity:(X ~ Obesity+Age)/ (X ~ Age) | **9.91E+03** | 1.11E-02 |
| Days since onset ~ Obesity+Age | **2.90E+03** | 2.49E-02 |
| Days since onset ~ Obesity | 2.95E-01 | 1.84E-05 |
| Effect of Obesity:(X ~ Obesity+Age)/ (X ~ Age) | **1.02E+04** | 1.10E-02 |
| Days since onset ~ Obesity+Age | 3.23E-01 | 1.04E-02 |
| Days since onset ~ Obesity | 2.65E-01 | 2.76E-05 |
| Effect of Obesity:(X ~ Obesity+Age)/ (X ~ Age) | 1.21E+00 | 2.86E-02 |

In the second column “model”, the effect of Obesity is made bold, because that is our most important model. In column “BF” any value for strong evidence (BF>10) for H_1_ is bold, any value indicative of strong evidence for H0 (no difference) is underlined. lmBF = Function to compute Bayes Factors for specific linear models. JZS = JeffreysZellner-Siow priors

**Supplementary Table 8.** Symptoms reported by C19+ participants with obesity (OB+) or without obesity (OB-).

| **Symptom** | **"Yes"** | | | | **"No"** | | | |  |  |
| --- | --- | --- | --- | --- | --- | --- | --- | --- | --- | --- |
|  | **OB+** | | **OB-** | | **OB+** | | **OB-** | |  |  |
|  | **n** | **prop** | **n** | **prop** | **n** | **prop** | **n** | **prop** | **Chi-square** | **P-value** |
| fever | 238 | 0.55 | 2190 | 0.52 | 195 | 0.45 | 2050 | 0.48 | 1.60 | 0.206 |
| dry cough | 243 | 0.56 | 2089 | 0.49 | 190 | 0.44 | 2151 | 0.51 | 7.11 | **0.008** |
| cough with mucus | 117 | 0.27 | 798 | 0.19 | 316 | 0.73 | 3442 | 0.81 | 16.26 | **0.000** |
| difficulty breathing/shortness of breath | 168 | 0.39 | 1352 | 0.32 | 265 | 0.61 | 2888 | 0.68 | 8.24 | **0.004** |
| chest tightness | 176 | 0.41 | 1308 | 0.31 | 257 | 0.59 | 2932 | 0.69 | 16.95 | **0.000** |
| runny nose | 177 | 0.41 | 1683 | 0.40 | 256 | 0.59 | 2557 | 0.60 | 0.18 | 0.669 |
| sore throat | 172 | 0.40 | 1535 | 0.36 | 261 | 0.60 | 2705 | 0.64 | 1.95 | 0.163 |
| changes in food flavor | 361 | 0.83 | 3488 | 0.82 | 72 | 0.17 | 752 | 0.18 | 0.26 | 0.610 |
| changes in smell | 386 | 0.89 | 3876 | 0.91 | 47 | 0.11 | 364 | 0.09 | 2.25 | 0.134 |
| loss of appetite | 225 | 0.52 | 1781 | 0.42 | 208 | 0.48 | 2459 | 0.58 | 15.50 | **0.000** |
| headache | 313 | 0.72 | 2994 | 0.71 | 120 | 0.28 | 1246 | 0.29 | 0.45 | 0.501 |
| muscle aches | 257 | 0.59 | 2376 | 0.56 | 176 | 0.41 | 1864 | 0.44 | 1.62 | 0.203 |
| fatigue | 338 | 0.78 | 3161 | 0.75 | 95 | 0.22 | 1079 | 0.25 | 2.39 | 0.122 |
| diarrhea | 195 | 0.45 | 1320 | 0.31 | 238 | 0.55 | 2920 | 0.69 | 34.03 | **0.000** |
| abdominal pain | 80 | 0.18 | 641 | 0.15 | 353 | 0.82 | 3599 | 0.85 | 3.14 | 0.076 |
| nausea | 114 | 0.26 | 907 | 0.21 | 319 | 0.74 | 3333 | 0.79 | 5.32 | **0.021** |

C19+ = positive COVID-19 diagnosis; OB+ = with obesity; OB+ = without obesity
